# SARS-CoV-2 infection induces the production of autoantibodies in severe COVID-19 patients in an age-dependent manner

**DOI:** 10.1101/2022.12.04.22282902

**Authors:** Dennyson Leandro M Fonseca, Igor Salerno Filgueiras, Alexandre HC Marques, Elroy Vojdani, Gilad Halpert, Yuri Ostrinski, Gabriela Crispim Baiocchi, Desirée Rodrigues Plaça, Paula P. Freire, Shahab Zaki Pour, Guido Moll, Rusan Catar, Yael Bublil Lavi, Jonathan I. Silverberg, Jason Zimmerman, Gustavo Cabral de Miranda, Robson F Carvalho, Taj Ali Khan, Harald Heidecke, Rodrigo JS Dalmolin, Andre Ducati Luchessi, Hans D. Ochs, Lena F. Schimke, Howard Amital, Gabriela Riemekasten, Israel Zyskind, Avi Z Rosenberg, Aristo Vojdani, Yehuda Shoenfeld, Otavio Cabral-Marques

## Abstract

Age is a significant risk factor for the coronavirus disease 2019 (COVID-19) outcomes due to immunosenescence and certain age-dependent medical conditions (e.g., obesity, cardiovascular disorder, diabetes, chronic respiratory disease). However, despite the well-known influence of age on autoantibody biology in health & disease, its impact on the risk of developing severe COVID-19 remains poorly explored. Here, we performed a cross-sectional study of autoantibodies directed against 58 targets associated with autoimmune diseases in 159 individuals with different COVID-19 outcomes (with 71 mild, 61 moderate, and 27 severe patients) and 73 healthy controls. We found that the natural production of autoantibodies increases with age and is exacerbated by SARS-CoV-2 infection, mostly in severe COVID-19 patients. Multivariate regression analysis showed that severe COVID-19 patients have a significant age-associated increase of autoantibody levels against 16 targets (e.g., amyloid β peptide, β catenin, cardiolipin, claudin, enteric nerve, fibulin, insulin receptor a, and platelet glycoprotein). Principal component analysis with spectrum decomposition based on these autoantibodies indicated an age-dependent stratification of severe COVID-19 patients. Random forest analysis ranked autoantibodies targeting cardiolipin, claudin, and platelet glycoprotein as the three most crucial autoantibodies for the stratification of severe elderly COVID-19 patients. Follow-up analysis using binomial regression found that anti-cardiolipin and anti-platelet glycoprotein autoantibodies indicated a significantly increased likelihood of developing a severe COVID-19 phenotype, presenting a synergistic effect on worsening COVID-19 outcomes. These findings provide new key insights to explain why elderly patients less favorable outcomes have than young individuals, suggesting new associations of distinct autoantibody levels with disease severity.

## INTRODUCTION

There is increasing evidence connecting coronavirus disease 2019 (COVID-19), caused by the severe acute respiratory syndrome virus 2 (SARS-CoV-2), with underlying autoimmune pathology^1,2^. The triggers of this intersection between COVID-19 and autoimmunity have been ascribed to both exacerbated and chronic inflammation^3^, e.g., by promoting the exposure to self-antigens and activation of bystander T cells caused by systemic high cytokine levels^4^, and due to the molecular mimicry between SARS-CoV-2 spike and human proteins^5–8^. Patients with severe COVID-19 develop profound organ damage due to a combination of several autoinflammatory and autoimmune responses, causing, among others, myopathy^9^, vasculitis, arthritis, antiphospholipid syndrome (APS)^10^ associated with deep vein thrombosis, pulmonary embolism, and stroke, as well as other organ damage to lungs, kidneys, and those forming the neurological system^11,12^. Furthermore, immune dysregulation is a hallmark of post-COVID syndrome^13^ causing heterogeneous symptoms such as fatigue, vascular dysfunction, pain syndromes, neurological manifestations, and neuropsychiatric syndromes^14–17^.

Following the initial discovery of autoantibodies against type I interferons (IFNs) in patients with life-threatening COVID-19^18^, several reports documented elevated levels of autoantibodies targeting various additional cytokines and chemokines and their receptors^19^, but also cardiac antigens^20^, G protein-coupled receptors (GPCR), renin-angiotensin system (RAS)-related molecules, and those against anti-cardiolipin^21–27^, ribosomal P proteins, chromatin proteins, thyroid antigens^28^, anti-nuclear antigen (ANA)^28,29^, and anti-neutrophil cytoplasmic proteins (ANCA)^30^ in patients with severe SARS-CoV-2 infections. We recently reported a large spectrum of autoantibodies linked to autoimmune diseases that associate with COVID-19 severity^31^. Autoantibody levels often accompany anti-SARS-CoV-2 antibody concentrations as essential predictors of COVID-19 outcome, together with age^31^.

Notably, aging has been strongly associated with increased morbidity and mortality of elderly patients with SARS-CoV-2 infections^32–34^. Elderly individuals present an increased risk of developing autoimmune diseases for several reasons, including immunosenescence and its associated immune dysregulation^35–37^, increased amounts of free DNA in the blood circulation^38^, and enhanced serum levels of autoantibodies^39,40^. Despite the well-known effect of age on autoantibody biology and immune pathophysiology in health & disease^1,3,11,41–43^, the particular influence of age in COVID-19 patients remains poorly explored. To address this issue, we performed a follow-up systems immunology analysis of our recent cross-sectional study of 159 individuals with different COVID-19 outcomes (mild, moderate, and severe) compared to 73 healthy controls^31,44^. We found that the natural production of autoantibodies increases with age and is exacerbated mainly in elderly patients with severe SARS-CoV-2 infections.

## METHODS

### Study cohort

We investigated 232 unvaccinated adults^44^, 159 COVID-19 patients with SARS-CoV-2 positive test by nasopharyngeal swab and polymerase chain reaction (PCR), and 73 randomly selected age- and sex-matched healthy controls who were SARS-CoV-2 negative and did not present any COVID-19 symptoms. COVID-19 patients were classified based on the World Health Organization (WHO) severity classification^45^ as mild COVID-19 (n=71; fever duration ≤ 1 day; peak temperature of 37.8 C), moderate COVID-19 (n=61; fever duration ≥ seven days; peak temperature of ≥ 38.8 C), and severe COVID-19 groups (n=27; severe symptoms and requiring supplemental oxygen therapy) (**Supplementary Table S1**). All healthy controls and patients provided informed written consent to participate in the study following the Declaration of Helsinki. The study was approved by the IntegReview institutional review board (Coronavirus Antibody Prevalence Study, CAPS-613) and followed the reporting guidelines of Strengthening the Reporting of Observational Studies in Epidemiology (STROBE).

### Measurements of anti-SARS-CoV-2 antibodies and autoantibodies linked to autoimmune diseases

Sera were assessed for the levels/titers of IgG anti-SARS-CoV-2 antibodies to spike and nucleocapsid proteins using the ZEUS SARS-CoV-2 ELISA Test System according to the manufacturer’s instructions (ZEUS Scientific, New Jersey, USA), as previously described^46^. We evaluated serum IgG autoantibodies against the nuclear antigen (ANA), extractable nuclear antigen (ENA), double-stranded DNA (dsDNA), actin, mitochondrial M2, and rheumatoid factor (RF) using commercial ELISA kits obtained from INOVA Diagnostics (San Diego, CA, USA). Furthermore, in a blinded fashion, we quantified IgG autoantibodies against 52 target molecules using an in-house ELISA procedure (Immunosciences Lab., Inc; Los Angeles, CA USA). One hundred mL of each autoantigen at the optimal concentration were prepared in 0.01 M PBS pH 7.4 and aliquoted into microtiter plates. We used a set of plates and coated each well with 2% bovine serum albumin (BSA) or human serum albumin (HSA) as controls. The ELISA plates were incubated overnight at 4°C and washed five times with 250 ml of 0.01 M PBS containing 0.05% Tween 20 pH 7.4. We avoided the non-specific binding of immunoglobins by adding 2% BSA in PBS and incubating the plates overnight at 4°C. The plates were washed, and the serum samples from healthy controls and SARS-CoV-2 patients were diluted 1:100 in serum diluent buffer or 1% BSA in PBS containing 0.05% Tween 20 and incubated for one hour at room temperature. The plates were rewashed, followed by the addition of alkaline phosphatase-conjugated goat anti-human IgG F(ab,)2 fragments (KPI, Gaithersburg, MD, USA) at an optimal dilution of 1:600 in 1% BSA PBS. The plates were incubated for an hour at room temperature and washed five times with PBS-Tween buffer. The enzyme reaction was started by adding 100 mL of para-nitrophenyl phosphate in 0.1 mL diethanolamine buffer 1 mg/mL plus 1 mM MgCl2 and sodium azide pH 9.8. Forty-five minutes later, the reaction was stopped with 50 mL of 1 N NaOH. The optical density (OD) was read at 405 nm using a microtiter plate reader. To exclude non-specific binding, the ODs of the control wells containing only HSA or BSA, always less than 0.15, were subtracted from those wells containing patient or control serum. The ELISA index for each autoantibody was calculated.

### Spearman’s correlation analysis

To evaluate the relationship between autoantibody levels, disease severity, and age, we used Spearman’s correlation analysis for the variables age and natural log of the sum of all autoantibodies and anti-SARS-CoV-2 antibody from healthy controls and each disease group (mild, moderate and severe COVID-19). All analyses were performed using R programming language version 4.2.1 (https://www.r-project.org/)^47^, and RStudio Version 2022.07.1+554^48^ with R package ggplot2^49^ for plots.

### Multivariable linear regression

To further explore the relationship between the variables age and specific autoantibody levels in each study group, we applied multivariable linear regression analysis^50^. This method evaluates the influence of age and study group on distinct antibody levels, thus, allowing to assess of the relationship between the levels of autoantibodies or the levels of anti-SARS-CoV-2 with age as a continuous variable of the study group (healthy control; mild, moderate, and severe COVID-19). Furthermore, patients’ sex was considered a covariable in the regression model since it represents a confounder that may influence the dependent variables. The formula used to calculate the crossing results is described below. We used the *lm* function from the R package stats for the linear regression analysis, and forest plots and scatter plots were generated using the R package ggplot2^51^. Autoantibodies as dependent variable (y); independent variables sex (a), age (b), and study group (c): y ∼ a+b*c.

### Differences in autoantibody levels and hierarchical clustering

We used box plots to show the distribution levels of autoantibodies in healthy controls and each COVID-19 group (mild, moderate, and severe), classifying individuals < 50 years of age as young and individuals ≥ 50 years of age as elderly (**Supplementary Table S1)**. Statistical differences in autoantibody levels were calculated using a two-sided Wilcoxon rank-sum test with a *p-value* < 0.05 as the significance cut-off. Box plots were generated using the R packages rstatix^52^, and ggplot2^51^. Additional visualization of autoantibody levels in the different study groups was performed using the R package ComplexHeatmap^53^ and Circlize^54^. The clustering of autoantibody levels in each study group was based on Euclidian distance.

### Principal Component Analysis

Based on the multivariable regression results, we identified significantly increased titers of age-associated autoantibodies against 16 targets, which underwent principal component analysis (PCA) with spectral decomposition^55,56^, as previously described^44,57^. This approach allowed us to measure the stratification power of the autoantibodies in distinguishing between severe COVID-19 patients and healthy controls while considering young and elderly groups. We calculated the eigenvalues based on the contributions of autoantibody levels to demonstrate their direction in the principal component analysis. The eigenvalues exceeding one intercept were considered essential to show the segregation of groups. For this, we used the R functions *get_eig* and *get_pca_var* from factoextra package^58^. PCA was performed using the function *prcomp* from the same package.

### Random forest modeling

We employed the random forest model to rank the most relevant autoantibodies (the autoantibodies that were significant in the multivariable regression analysis) to best classify COVID-19 disease severity for each age category (young and elderly) using the R package randomForest (version 4.7.1.1)^59^ as previously described^31,44,60^. Briefly, five thousand trees were used, and three variables were resampled (mtry parameter). As criteria to determine variable importance in the classification, we considered the mean minimum depth, Gini decrease, and the number of appearances in nodes. The dataset was split into training and testing sets using a 3 to 1 ratio for cross-validation, while quality was assessed for each, respectively, through out-of-bags error rate and the ROC curve.

### Support Vector Machine (SVM) classification

We used support vector machine^61,62^ (SVM), a robust computer algorithm, to build classifiers^63^. SVM employs four basic concepts: separating hyperplane, the maximum-margin hyperplane, the soft margin, and the kernel function^64^. We performed the radial kernel function applied between healthy controls and the severe COVID-19 group to classify the scaled values of the anti-cardiolipin and anti-platelet glycoprotein autoantibodies with age. Groups were defined as the dependent variable, while antibodies and age were considered independent variables. The analysis was performed using the *svm* function of the e1071^65^ R package. We used the kernel (C-classification) with 50% of our data sorted randomly by the R base *sample* function for training and predicting, considering the *radial basis* parameter: *exp(-gamma\**|*u-v*|*^2)*, which was the best model applied to our data. Accuracy was defined as the percentage of correctly classified samples resulting in 77% for cardiolipin and 81% for platelet glycoprotein, correctly classified as healthy controls and severe COVID-19 patients in our model. Furthermore, we used the *tune* function of the R package e1071^65^ to adjust the hyperparameters for cost and gamma in the *svm* function. We used a cost of 10 and a gamma of 0.5 for our data. All graphs resulting from *svm* prediction results were generated using the R package ggplot2^51^.

### Binomial logistic regression

We used the binomial logistic regression analysis to understand whether the severity of COVID-19 (dichotomous dependent variable) can be predicted based on age and autoantibody levels (independent variables). The binomial logistic regression analysis indicates the probability that an observation falls in one of two defined dichotomous categories (dependent variable) based on one or more independent variables (either continuous or categorical)^66^. This analysis was performed using the R package stats^47^ with the *glm* function. The categories of the dichotomous dependent variable were defined as “belonging to severe COVID-19: group 1” and “not belonging to severe COVID-19: group 0”, using the binomial logistic family to predict the probability of falling into the severe COVID-19 group in relation to the healthy controls. The age categories, young and elderly, and the levels of autoantibodies targeting cardiolipin, platelet glycoprotein, and claudin-5 were the independent variables for this analysis. This approach resulted in a regression coefficient and *p-value* for the probability of severe COVID-19 based on the autoantibody level and the likelihood of severe COVID-19 based on the age category.

### Linear discriminant analysis

We applied linear discriminant analysis (LDA), a method to find a linear combination of variables that characterize or separate two or more classes of objects or events^67^, to classify in which group (elderly healthy controls: group 0; elderly severe COVID-19: group 1) the sample elements (significant age-associated autoantibodies targeting 16 molecules) belong to. Autoantibodies with a specificity and sensitivity value greater than 70% (anti-cardiolipin and anti-platelet glycoprotein) were considered to be related to the elderly severe COVID-19 group. The analysis was performed using the R package MASS^68^ with the *lda* function. To plot the specificity and sensitivity of the class prediction for each autoantibody, we used the R package plotROC^69^ and ggplot2^51^. In addition, we performed the binomial logistic regression for odds ratio (OR)^66^ as described above using autoantibodies with specificity and sensitivity greater than 70%. The calculation of the chance to predict COVID-19 severity was performed using the function *logistic*.*display* of the R package epiDisplay^70^. Here we demonstrate the odds ratio when there is an increase in cardiolipin or platelet glycoprotein levels alone or combined. Plots resulting from this analysis were generated using the R package forestploter^71^ and the *forest* and *forest_theme* functions.

## RESULTS

### Autoantibody levels increase with age, particularly in severe COVID-19 patients

We observed that, in general, the autoantibody levels increased by age across all groups included in this study. However, autoantibody levels in severe COVID-19 patients presented the strongest correlation with age (ρ = 0,62 and *p* = 0,00019; **Figure 1A**) and the higher mean levels (>1 Units/mL; **Figure 1C**) when compared to healthy individuals, as well as mild and moderate COVID-19 groups. Interestingly, these results contrast with the levels of anti-SARS-CoV-2 antibodies, which showed no correlation with age at all for all tested groups (ρ *< 0*.*0* and *p* = 0.3 to 0.7; **Figures 1B**) but generally increased with disease severity (from 1-5 to 2 Units/mL; **Figure 1D**). Details are available in the **supplementary Table S2**.

**Figure 1.**
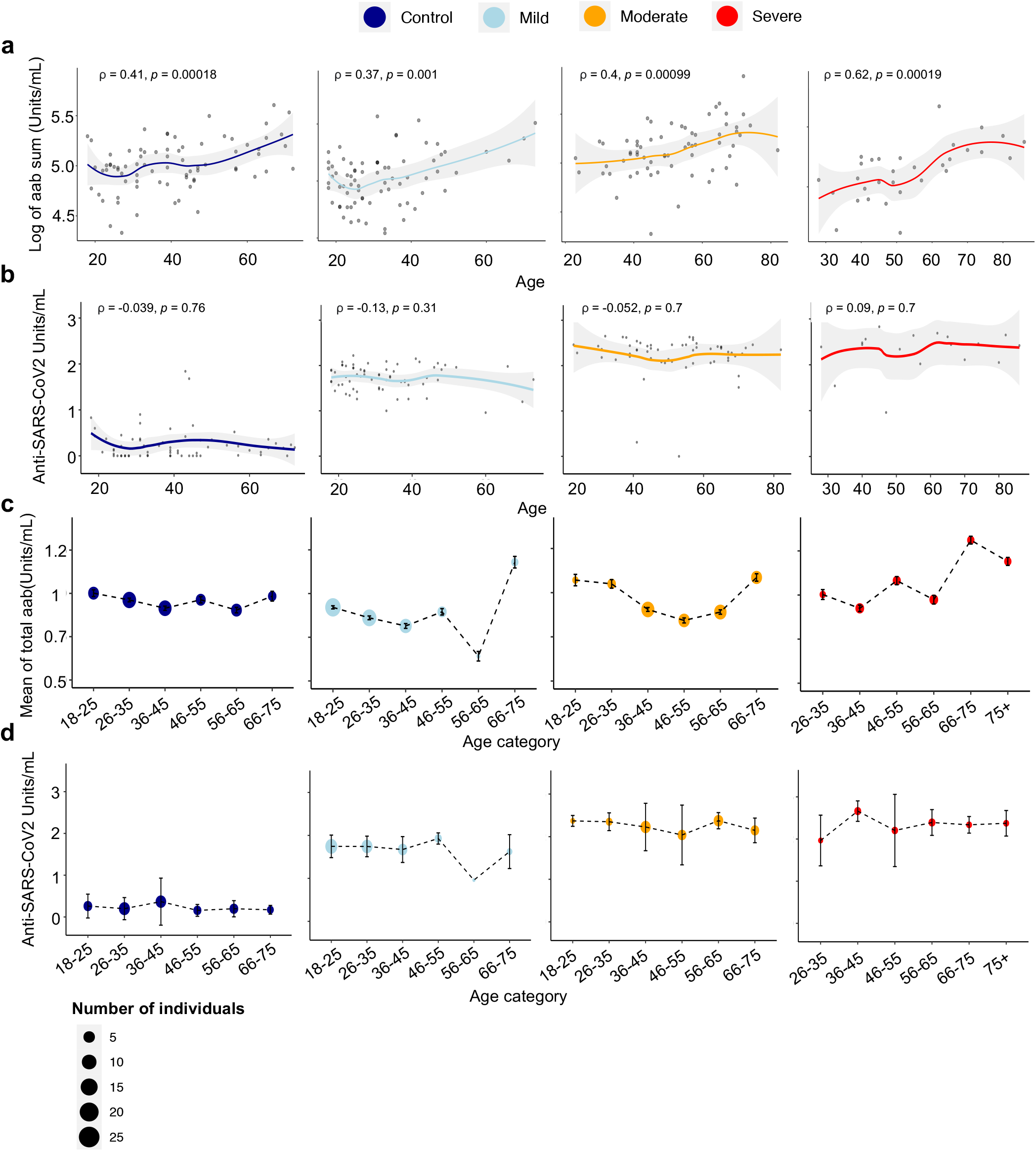
Increased autoantibody levels by age. **a-b**) Scatter plot indicating the relationship and correlation between age and (**a**) the natural log of the sum of autoantibodies targeting 52 different molecules and (**b**) anti-SARS-CoV-2 levels in healthy controls and each COVID-19 disease group. Spearman’s rank correlation coefficient (⍰) and significance level (*p*-value) for the correlations are shown within each graph. Healthy controls n=73; COVID-19 groups: mild n=71, moderate n=61, and severe n=27. **c-d**) Graphics showing the relationship between the mean of **c**) autoantibodies and **d**) anti-SARS-CoV-2 levels in different age categories for healthy controls and COVID-19 disease groups. The size of the dots corresponds to the number of individuals in the age category according to the figure legend (see **Supplementary Tables S1** and **S2)**.

To characterize which autoantibodies significantly contributed to the age-associated enhancement in the autoantibody levels, we performed a linear regression analysis for each autoantibody. In this context, autoantibodies were considered the dependent variable, while group and age were evaluated as independent variables. In agreement with the descriptive statistical analysis shown in **Figure 1**, this inferential approach revealed autoantibodies targeting sixteen molecules, which were strongly significantly associated with age in the severe COVID-19 group compared with the healthy controls (**Figure 2A**). In contrast, except for the considerable significant enhancement in the levels of autoantibodies targeting claudin 5 and transglutaminase 6 in the mild COVID-19 group, there was only a general non-significant trend for increasing autoantibody levels for the mild and moderate COVID-19 groups (**Figures 2A** and **2B; Supplementary Table S3**; see **Supplementary Figure 1** for the comparison of all groups). This indicates that many autoantibody levels robustly increase with age, particularly in severe COVID-19 patients, but lesser in the mild and moderate COVID-19 patients

**Figure 2.**
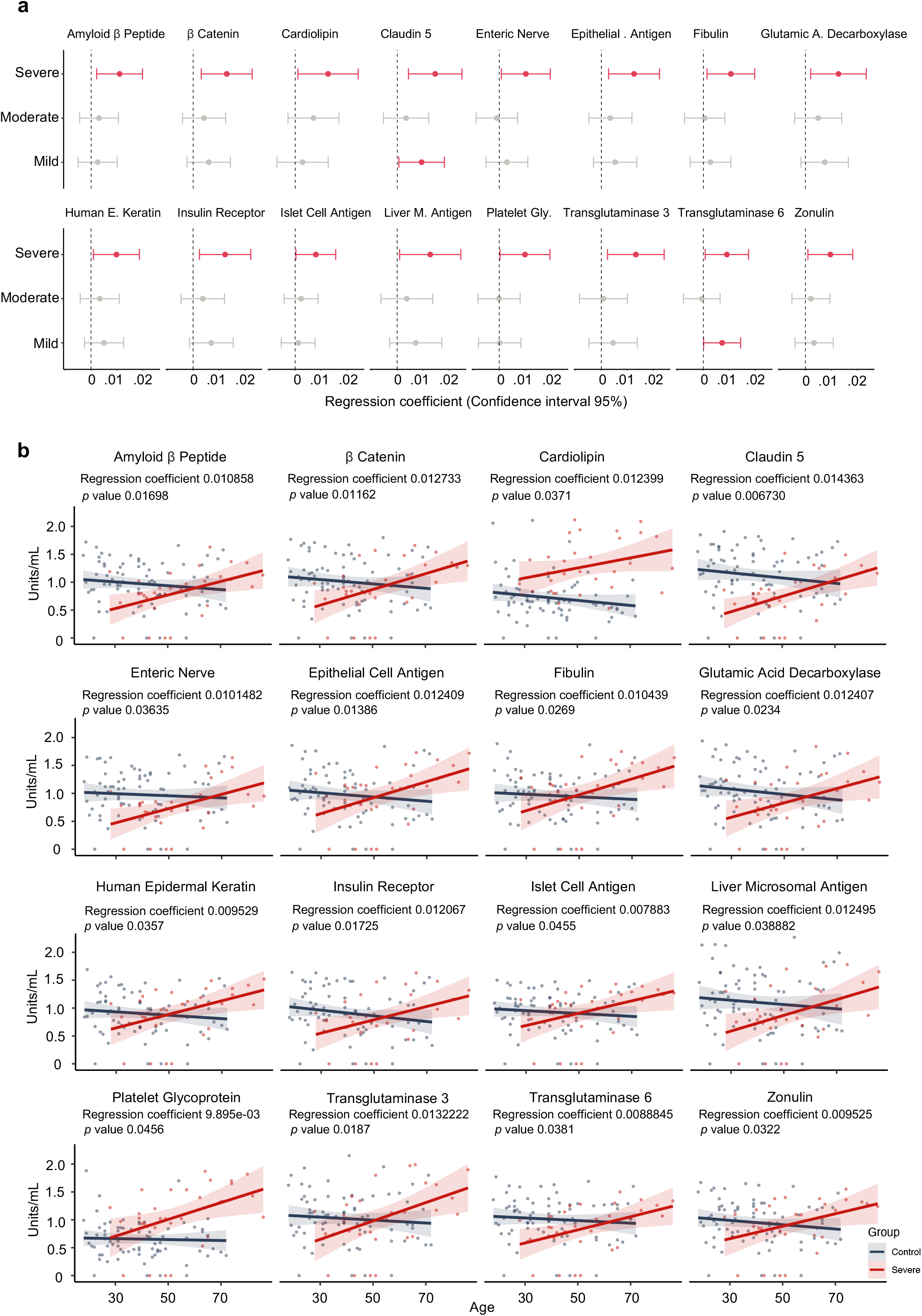
Regression analysis demonstrating the relationship between autoantibodies and age in severe COVID-19 patients. **a)** Forest plots showing linear regression coefficients (dots) and their 95% confidence interval (whiskers) for different autoantibodies across the COVID-19 groups (mild, moderate, and severe) compared to healthy controls (vertical dotted line at the intercept of 0). Red dots and lines correspond to significantly increased autoantibody levels associated with disease group and age compared to healthy controls. **b)** Scatter plot of regression analysis, indicating the relationship between the autoantibodies and age for severe COVID-19 and control groups. The *p*-values and linear regression coefficient are indicated for each graph. Scatter plots for mild and moderate patient groups are shown in **Supplementary Figure 1. Supplementary Table S3** shows the results of all regression coefficients.

### Hierarchical clustering of autoantibodies by age indicates the segregation of young from elderly severe COVID-19 patients

To further investigate the impact of age on the levels of autoantibodies, we divided the healthy controls and the COVID-19 patients by age, i.e., individuals <50 years old as the young subgroup and those ≥50 years old as the elderly subgroup for each category (healthy controls as well as mild, moderate, and severe COVID-19 patients). In agreement with the continuous autoantibody values analyzed by the linear regression analysis (**Figure 2**), this approach revealed that elderly healthy controls and all elderly COVID-19 groups tended to have higher autoantibody levels compared to their young counterparts. However, only the severe elderly COVID-19 patients showed many significantly higher levels of autoantibodies targeting various autoantigens (e.g., platelet glycoprotein, amyloid β peptide, and β catenin) when compared to the severe young COVID-19 (**Figure 3a; Supplementary Table S4**). Likewise, hierarchical clustering analysis of the autoantibody levels based on the aging groups (young versus elderly individuals) uncovered a segregation of young from elderly severe COVID-19 patients but not the other groups investigated (**Figure 3b**). This suggests a synergistic effect of patient age and COVID-19 severity influencing the positive association among the autoantibodies.

**Figure 3.**
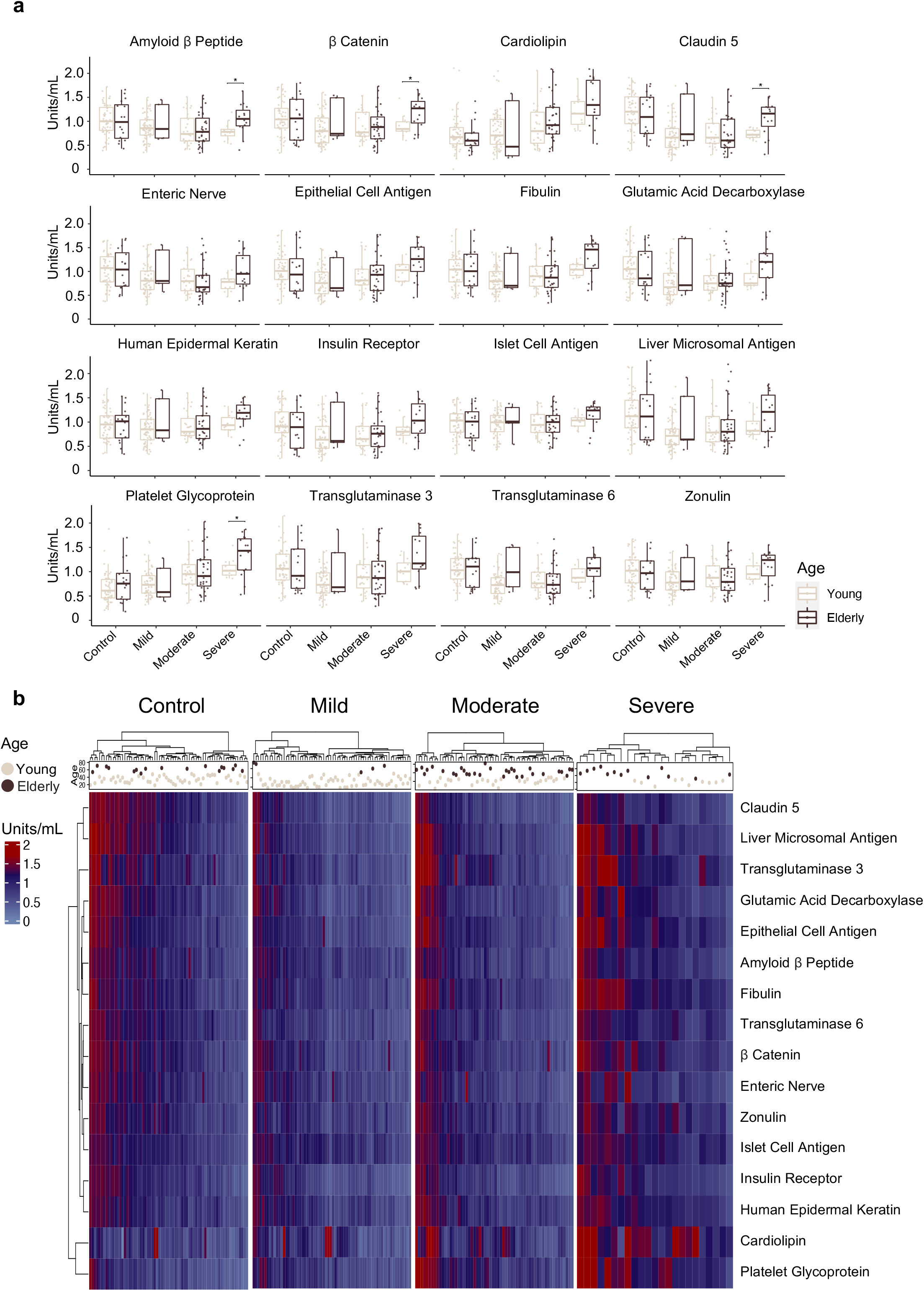
Autoantibody levels are increased in elderly severe COVID-19 patients. **a)** Boxplots showing the autoantibody levels in young (< 50 years old) and elderly (≥ 50 years old) groups for healthy controls as well as mild, moderate, and severe COVID-19 patients. The difference in autoantibody levels comparing young with elderly individuals of each group was calculated using the nonparametric Wilcoxon test considering a *p-value* < 0.05 as significant (denoted by * in the plot). See **Supplementary Table S4** for the exact numbers of *p*-values. **b)** Heatmaps showing autoantibody levels ranging from 0 to 2 Units/ml according to the color scale bar at the side of the graph clustered by Euclidian Distance for each disease and control group. The age and age categories (light grey and brown dots above the heatmap for the young and elderly categories, respectively) for all individuals are shown above the heatmap.

### Autoantibodies associated with age stratify COVID-19 patients

Next, we performed random forest analysis to rank the most critical autoantibodies linked to severe COVID-19 and aging among the sixteen age-associated autoantibodies. This approach identified autoantibodies targeting claudin 5, cardiolipin, and platelet glycoprotein as the three most essential autoantibodies classifying young severe COVID-19 versus young, healthy controls and elderly severe COVID-19 versus elderly healthy controls (**Figure 4a**). The receiver operating characteristic (ROC) curves of these comparisons demonstrate the high accuracy of the random forest analysis based on the age-associated autoantibodies as classifiers of severe COVID-19 patients (**Figures 4b-4c and Supplementary Table S5**).

**Figure 4.**
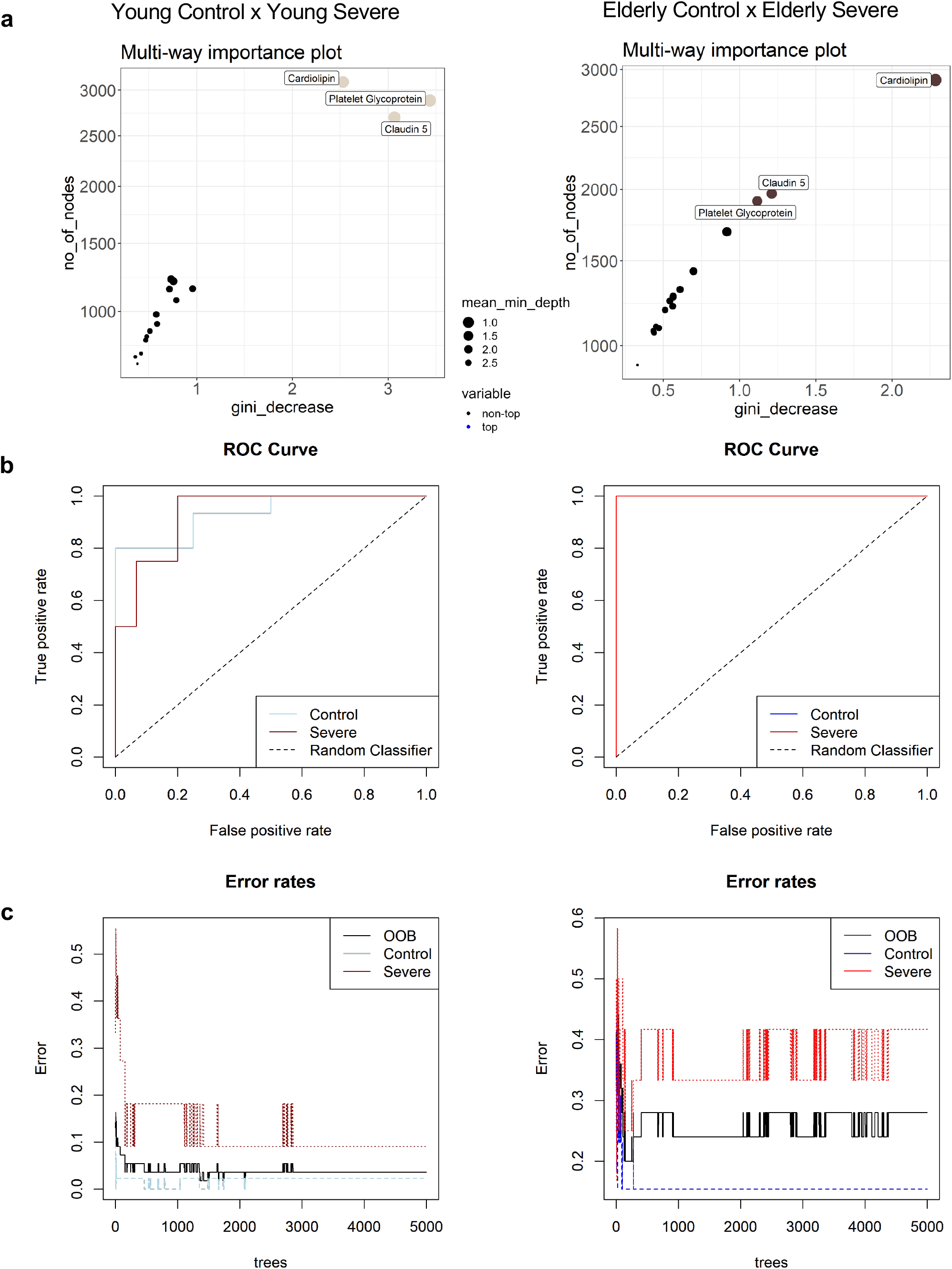
Ranking of autoantibodies as predictors for severe COVID-19 in young and elderly groups. **a)** Random Forest model used to rank the 16 most significant autoantibodies as predictors for severe COVID-19 in young and elderly severe COVID-19 patient groups compared to healthy controls. Multi-way importance plots show four nodes (IgG antibodies), the most significant predictors of severe COVID-19 in the young and the elderly. The size of the dots corresponds to the mean min depth in decreasing order (2.5 to 1.0) **b)** ROC curve of the random forest model showing over 70% accuracy in specificity and sensitivity for healthy controls and severe COVID-19 groups of each age category (young and elderly). **c)** Stable curve showing the number of trees and out-of-bag (OOB) error rate of 20% for young healthy controls and 9% for the young severe COVID-19 group (left side graph of the figure), as well as an error rate of 15% for elderly healthy controls and 41% for the elderly severe COVID-19 group (right side graph of the figure). **Supplementary Table S5** shows the confusion matrix for the random forest model.

To further investigate potentially age-related autoantibodies that stratify young from elderly severe COVID-19 patients and healthy controls, we carried out PCA based spectral decomposition^72^. The PCA showed that while young and elderly healthy controls presented a similar autoantibody pattern, there is a stratification of elderly and young severe COVID-19 patients from elderly and young healthy controls (**Figure 5A**). According to eigenvalue criteria, this may be viewed for just two dimensions (Intercept > 1; **Figure 5B**). Although not at the same stratification magnitude when comparing healthy controls with severe COVID-19 patients, the PCA suggested segregation between elderly and young severe COVID-19 patients, shown by dimension 2 in the y-axis (**Figure 5A**). In agreement with the random forest results, autoantibodies targeting claudin 5, cardiolipin, and platelet glycoprotein mainly contributed to dimension 2, which was responsible for the stratification of the control and severe groups (**Figures 5A** and **5C**; **Supplementary Tables S6** and **S7**).

**Figure 5.**
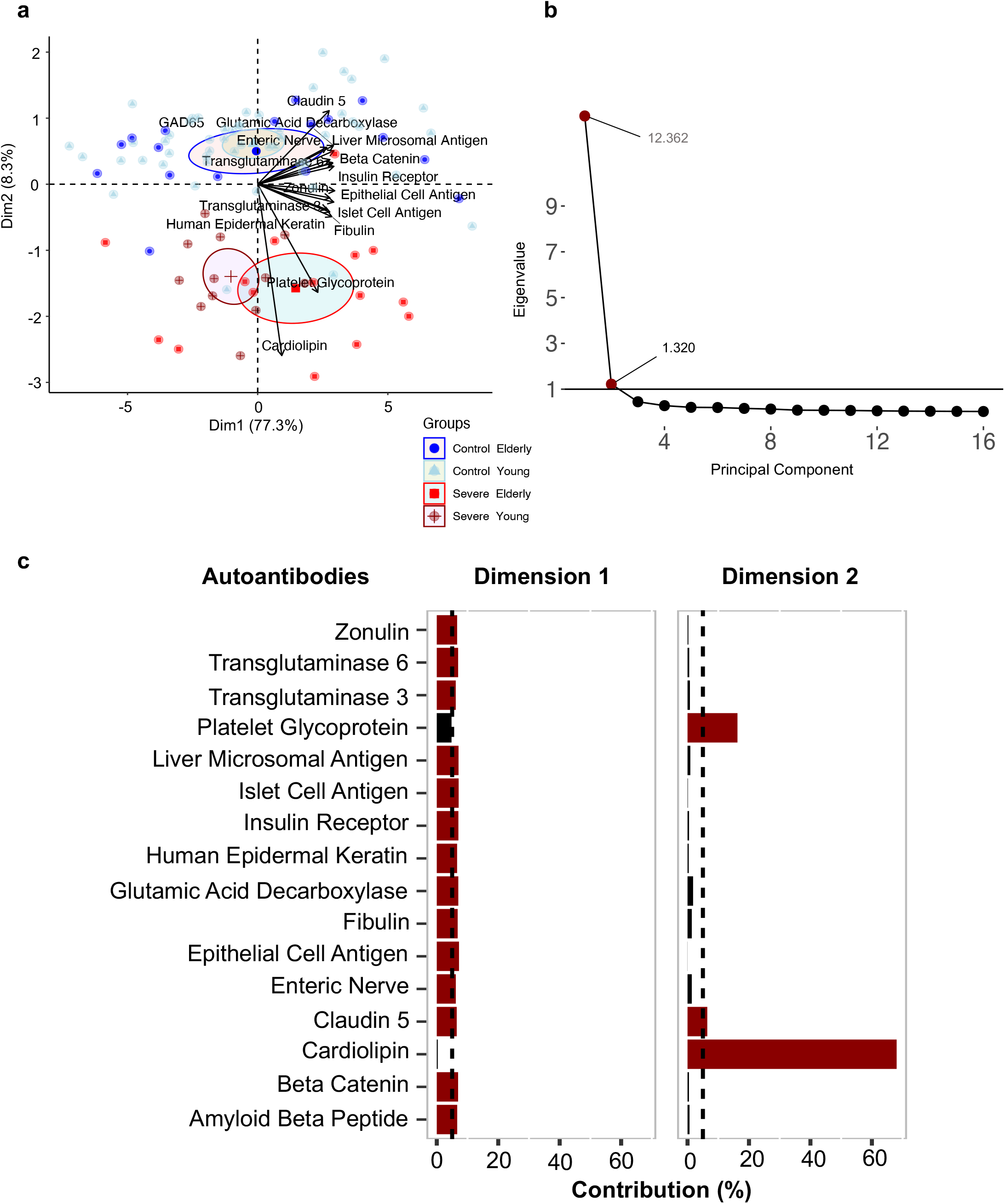
A subset of autoantibodies differentiates between severe COVID-19 and healthy controls in young and elderly age categories. **a)** Principal component analysis (PCA) with spectral decomposition shows the stratification power of the sixteen most significant autoantibodies to distinguish between severe COVID-19 and healthy controls, considering the age categories of each group according to the first and second dimensions. **b)** red dots show eigenvalues above one, and eigenvalues below 1 are shown by black dots demonstrating the importance of the dimensions (principal component). The horizontal black line shows the intercept of 1. Eigenvalues are available in **Supplementary table S6. c)** Barplots for two dimensions based on variable contribution. Each barplot shows the contribution (in %) of the sixteen autoantibodies to each dimension. The red colored bars represent contribution values ≥ 5% (black dashed intercept line), while black colored bars indicate contribution values < 5%. The contribution values of all autoantibodies to the different dimension is listed in **Supplementary Table S7**.

### Age-associated autoantibodies increase the probability of disease severity

The results above have suggested the importance of the autoantibodies targeting claudin 5, cardiolipin, and platelet glycoprotein in the classification of COVID-19 patients. We conducted a multivariate logistic regression analysis to understand better their contribution to developing severe COVID-19 disease. This approach was used to understand the relationship between the groups (healthy controls versus COVID-19 groups) as the dependent variable and the autoantibody levels as the independent variable to predict the likelihood of COVID-19 severity. This approach indicated a significant probability of developing severe COVID-19 directly proportional to the levels of anti-cardiolipin (*p*=8,92e-06) and anti-platelet glycoprotein (*p*= 0.001) autoantibodies (**Figure 6A)**. In particular, the severe elderly COVID-19 patients exhibited a higher probability curve of disease severity when compared to severe young COVID-19 patients (**Figure 6A**) and increased levels of anti-cardiolipin and anti-platelet glycoprotein (*p* = 0.01 to *p* = 0.001; **Figure 6B**; **Supplementary Table S8**). Conversely, the levels of autoantibodies targeting claudin 5 presented a significant inversely proportional relationship with the probability of developing severe COVID-19 in young (*p*<0.01; **Figure 6B**) but not in elderly patients.

**Figure 6.**
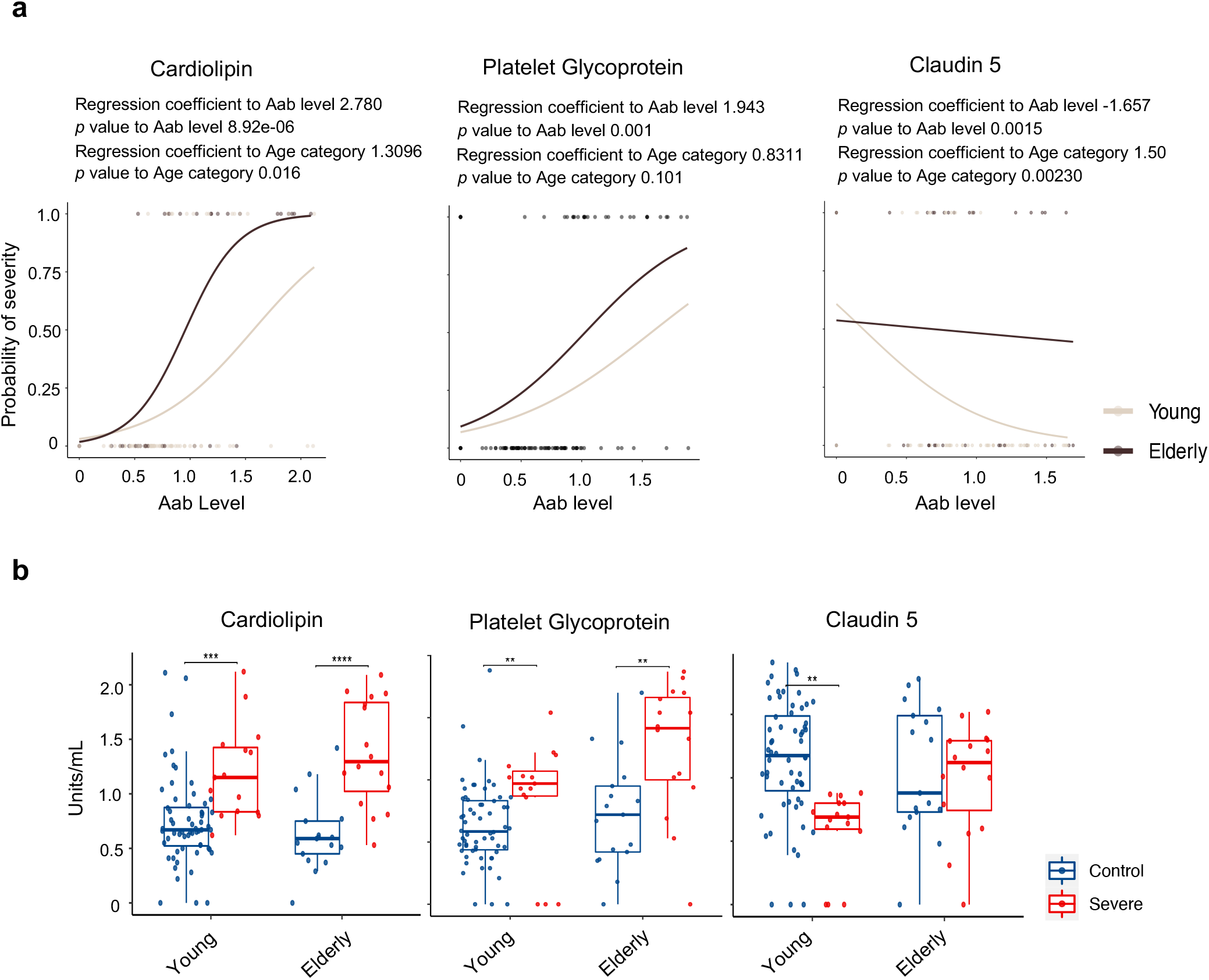
The probability of disease severity associated with autoantibody levels. **a)** Scatter plot of three autoantibodies (anti-Cardiolipin, anti-Platelet Glycoprotein, and anti-Claudin 5) show that increased autoantibody levels can be explained by a higher probability of being severe in each group (healthy controls = 0, and severe COVID-19 = 1). The age category for each group (healthy controls and severe COVID-19) is indicated by a light grey (young) and brown (elderly) line in the center of the graph. The regression coefficients to autoantibody levels and comparisons between young and elderly age categories are shown above each graph. **b)** Boxplots indicate the autoantibody levels of the three autoantibodies (Cardiolipin, Platelet Glycoprotein, and Claudin 5) in healthy controls and severe COVID-19 patients for young and elderly categories. Significant differences were defined using the nonparametric Wilcoxon test and *p-value* < 0.05 (denoted by * in the plot). See **Supplementary Table S8** for the exact number of *p*-values.

Of note, the support vector machine classification, which is a powerful machine learning approach with maximization (support) of separating margin (vector)^63,64^, based on the levels of anti-cardiolipin (**Figure 7a**) or anti-platelet glycoprotein (**Figure 7b**) in relation to age, showed the importance of these autoantibodies as suitable classifiers of severe COVID-19 when compared to healthy controls. i.e., SVM showed the separation of severe COVID-19 from healthy controls based on these most critical age-associated autoantibodies as the random forest analysis predicted. The input data and table results are shown in **Supplementary Figure 2** and **Supplementary Tables S9** and **S10**. We next used an LDA model considering the groups (elderly healthy controls versus elderly severe COVID-19 patients) as the dependent variables and the autoantibody levels as the independent variable to calculate the OR of disease severity (**Supplementary Tables S11** and **S12**). However, the binominal logistic regression analysis indicated only anti-cardiolipin and anti-platelet glycoprotein (when considering the sixteen aged-associated autoantibodies) with specificity, sensitivity, and accuracy above 70% chance of correct group classification (**Figure 7c** and **Supplementary Figure 3**). The OR of disease severity indicated that autoantibodies targeting cardiolipin or platelet glycoprotein highly increase the chance of developing severe COVID-19. Notably, the odds of developing severe COVID-19 at least double when the levels of these two autoantibodies increase (**Figure 7d**), thus indicating their possible relevance in the pathophysiology of severe COVID-19

**Figure 7.**
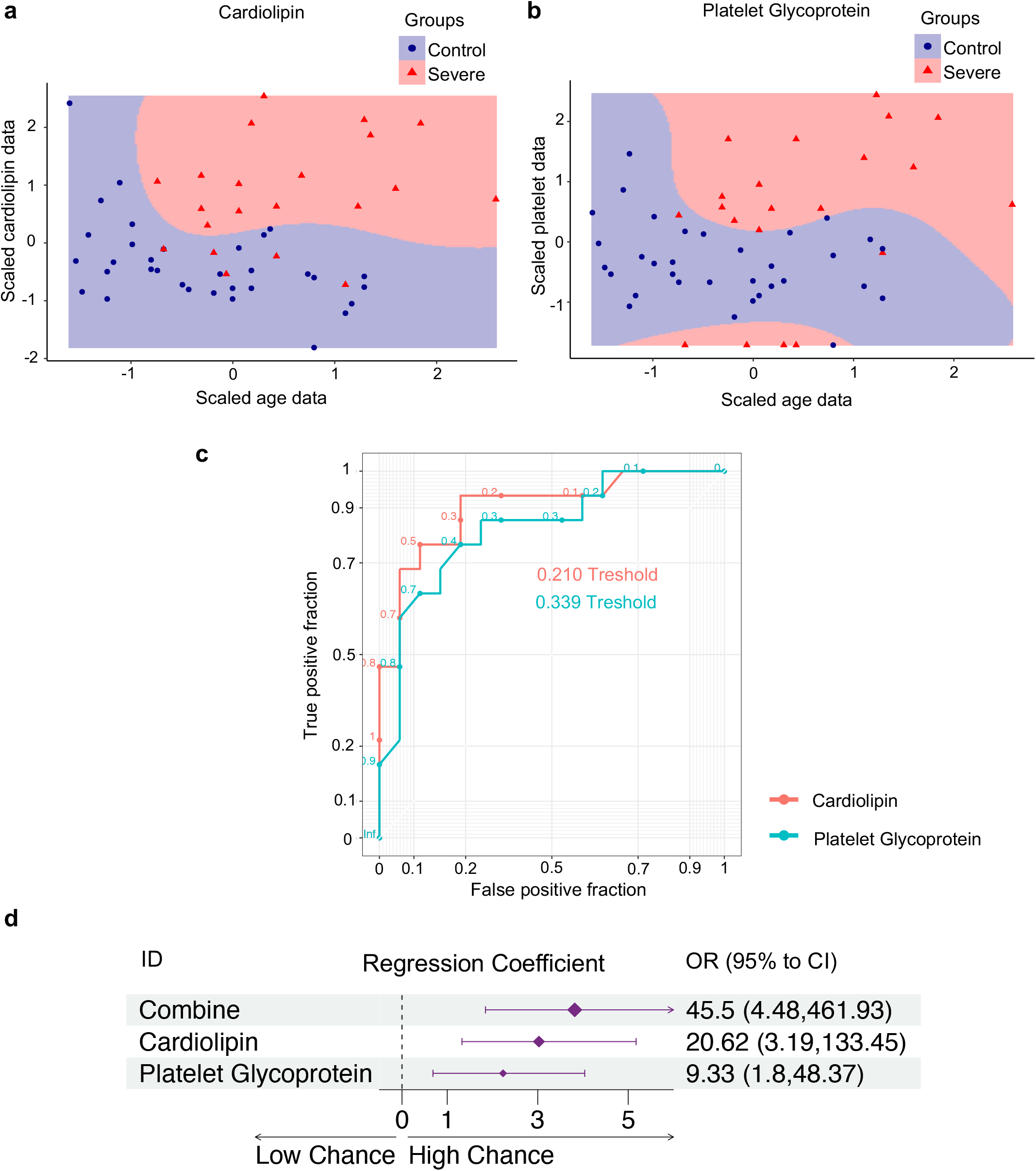
Increased levels of anti-cardiolipin and anti-platelet glycoprotein autoantibodies indicate a higher chance of severity in the elderly. **a-b)** Support Vector Machine (SVM) showing the non-linearly C-classification based on radial kernel with over 70% (**Supplementary Figure 2 and Supplementary Table S10**) accuracy between healthy controls and the severe COVID-19 group of the elderly age category. The scaled values for age (x-axis) and anti-cardiolipin, and (**a**) anti-platelet glycoprotein (b) autoantibody levels (y-axis) are shown. The colors indicate each study group according to the figure legend. **c)** ROC curve and threshold values for the two autoantibodies based on linear discriminant analysis (LDA) representing the specificity and sensitivity of each autoantibody for severe COVID-19 in the elderly category (**Supplementary Figure 3** shows the ROC curve of all sixteen autoantibodies). **d)** Forest plot with Odds radial (OR) from binomial logistic regression analysis showing the regression coefficient (dots) with confidence intervals (whiskers) and the significance level (≠ 0 intercept). The exact values of OR and 95% confidence intervals are shown on the side of the regression coefficients.

## DISCUSSION

Here we show the distinct impact of patient aging on the level of serum autoantibodies and COVID-19 severity, which we previously found to be associated with autoimmune diseases in patients with COVID-19^44,73,74^. Increasing evidence has indicated that SARS-CoV-2 infection triggers a life-threatening immune dysregulation, with the recent demonstration of autoantibody production targeting an array of self-antigens^19,28,75,76^. However, the effect of patient aging in the context of autoimmune homeostasis was not considered in these recent studies investigating the phenomenon of autoreactivity in COVID-19 patients^77^.In line with the already well-known impact of patient aging, which is one of the most decisive risk factors for the development of severe COVID-19^78^ (in addition to other strong risk factors, such as obesity and prehistory of cardiovascular complications), our new data suggest that the production of natural autoantibodies (but not anti-SARS-CoV-2 antibodies) is significantly increased in an age-dependent manner, being most pronounced in individuals with severe COVID-19.

Hierarchical clustering analysis of autoantibody levels indicated a segregation of young from elderly patients with severe COVID-19. Notably, the combination of different machine learning approaches revealed that, among the significantly age-associated autoantibodies, particularly those directed against cardiolipin and platelet glycoprotein, are the most critical autoantibodies for predicting severe elderly COVID-19 patients when compared to elderly healthy controls.

Of note, linear multivariable and binominal logistic regression analyses indicated that autoantibodies targeting cardiolipin and platelet glycoprotein synergistically increase the probability of developing severe disease. Thus, in addition to the impaired immune response (affecting IFN-mediated immunity) and the generation of anti-type I IFN autoantibodies that drive the age-dependent severity of COVID-19^79,80^, patients with life-threatening SARS-CoV-2 infections also present with an age-dependent increase of multiple autoantibodies associated with classic autoimmune diseases that correlate with disease severity.

The well-documented observation that anti-cardiolipin^81^ and anti-platelet antibodies^82^ increase the risk of thrombosis-related events such as pulmonary thromboembolism and deep vein thrombosis^83,84^ is also true for COVID-19 patients^11,21,24,85–87^. Thus, our findings could provide new insights into the complex pathophysiology of COVID-19, such as the thrombosis-related pathological events occurring with increased frequency in elderly individuals with SARS-CoV-2 infection. However, this represents a limitation of our study since we have no longitudinal data of our patients to evaluate if the individuals with high levels of anti-cardiolipin and anti-platelet antibodies developed thrombosis-related events.

The elevated autoantibody levels associated with severe COVID-19 may be exacerbated by the evolutionarily conserved tendency to produce more autoantibodies with increasing age^42^. This phenomenon can aggravate the age-associated deficit in cardiovascular structure and function^88^ as well as the age-related decline of normal lung function^89^, which represent two central physiological systems (circulatory and respiratory) that are predominantly harmed in COVID-19 patients^90^. Together, these age-associated conditions create a fertile milieu for the poor outcomes of elderly individuals suffering from severe SARS-CoV-2 infection.

Our results raise the important question, considering the sequence of the underlying events: does the severity of COVID-19 increase the autoantibody levels? Or do the increased autoantibody levels affect the disease severity? We here hypothesize that both possibilities are reasonable and may be complementary. The severe COVID-19 infection promotes a body environment, i.e., the SARS-CoV-2 induced immune dysregulation, which is favorable for the production of autoantibodies, which could act synergistically with multiple metabolites^91^, cytokines, and chemokines, which are naturally dysregulated in elderly patients as part of immunosenescence^35–37^, worsening the COVID-19 outcomes through several well-known mechanisms of autoantibody-induced pathology^92^ In this context, autoantibodies - in concert with other immune molecules (e.g., cytokines and chemokines) - could interact in a highly complex network underlying immunopathological processes^93^ in severe COVID-19 patients, potentiated by aging-associated health conditions and lead to the development of severe disease.

Another age-dependent phenomenon that possibly explains the increased autoimmune responses we observed in the elderly patients with severe COVID-19 relies on the accumulation of epigenetic alterations (e.g., DNA methylation and histone acetylation)^94^, known to contribute to the autoimmunity risk of elderly individuals. Accordingly, accelerated epigenetic aging has been associated with the increased risk of SARS-CoV-2 infection and the development of severe COVID-19^95^. Lastly, a state of hyper-stimulation of the immune system by the SARS-COV-2 infection has been observed in elderly patients, for instance, by promoting the activation of overlapping B cell pathways between severe COVID-19 and patients with systemic autoimmune diseases^96^. Hence, several age-associated immunopathological events support the existence of age-associated autoantibodies, increasing the likelihood of severe COVID-19 disease in elderly patients.

Importantly, our data indicate a distinct separation/stratification of the young from the elderly COVID-19 patients and an increased odds ratio of disease severity due to high levels of autoantibodies known to be associated with classic autoimmune diseases, in particular those targeting cardiolipin or platelet glycoprotein. Indeed, the prothrombotic anti-cardiolipin autoantibodies that may potentially exacerbate the thrombo-inflammatory state related to severe COVID-19^21,97^, and other autoantibodies linked to classic autoimmune diseases^31^, have long been known to be highly prevalent in the healthy elderly population^98,99^.

Since we were unable to measure the autoantibody levels of our patient cohort before the SARS-CoV-2 infection (which is one of the fundamental constraints of many studies), we cannot reject the possibility that at least some of our patients already had elevated levels of age-associated autoantibodies before the development of severe COVID-19. Thus, our findings could be influenced by a predisposition factor for the association between age, autoantibody levels, and severe COVID-19. Therefore, our results require future mechanistic investigations through *in vivo* approaches, as recently suggested^19,79^.

In conclusion, our data provide new crucial insights into the critical relationship between severe COVID-19 and the increased dysregulation/production of distinct autoantibodies with increasing age that may be an essential component associated with the development of severe COVID-19. As demonstrated by the stratification of young from elderly COVID-19 patients and the increased odds ratio of disease severity due to the high levels of autoantibodies linked to autoimmune diseases, in particular, those targeting cardiolipin or platelet glycoprotein, our data indicate an age-dependent effect of autoantibodies in the development of severe COVID-19, that may also be of future value for disease prognosis. This work expands the link between senescence and aging with severe SARS-CoV-2 infection^100–105^.

## Supporting information

Raw data of study cohort

## Data Availability

All data produced in the present work are contained in the supplementary tables

## Acknowledgments

We thank the São Paulo Research Foundation (FAPESP grants 2018/18886-9, 2020/01688-0, and 2020/07069-0 to OCM, 2020/16246-2 to DLMF, 2020/09146-1 to PPF, 2020/07972-1 to GCB, 2020/11710-2 to DRP) for financial support. FAPESP and CAPES supported computational analysis.

## SUPPLEMENTARY FIGURES

**Supplementary Figure 1.**
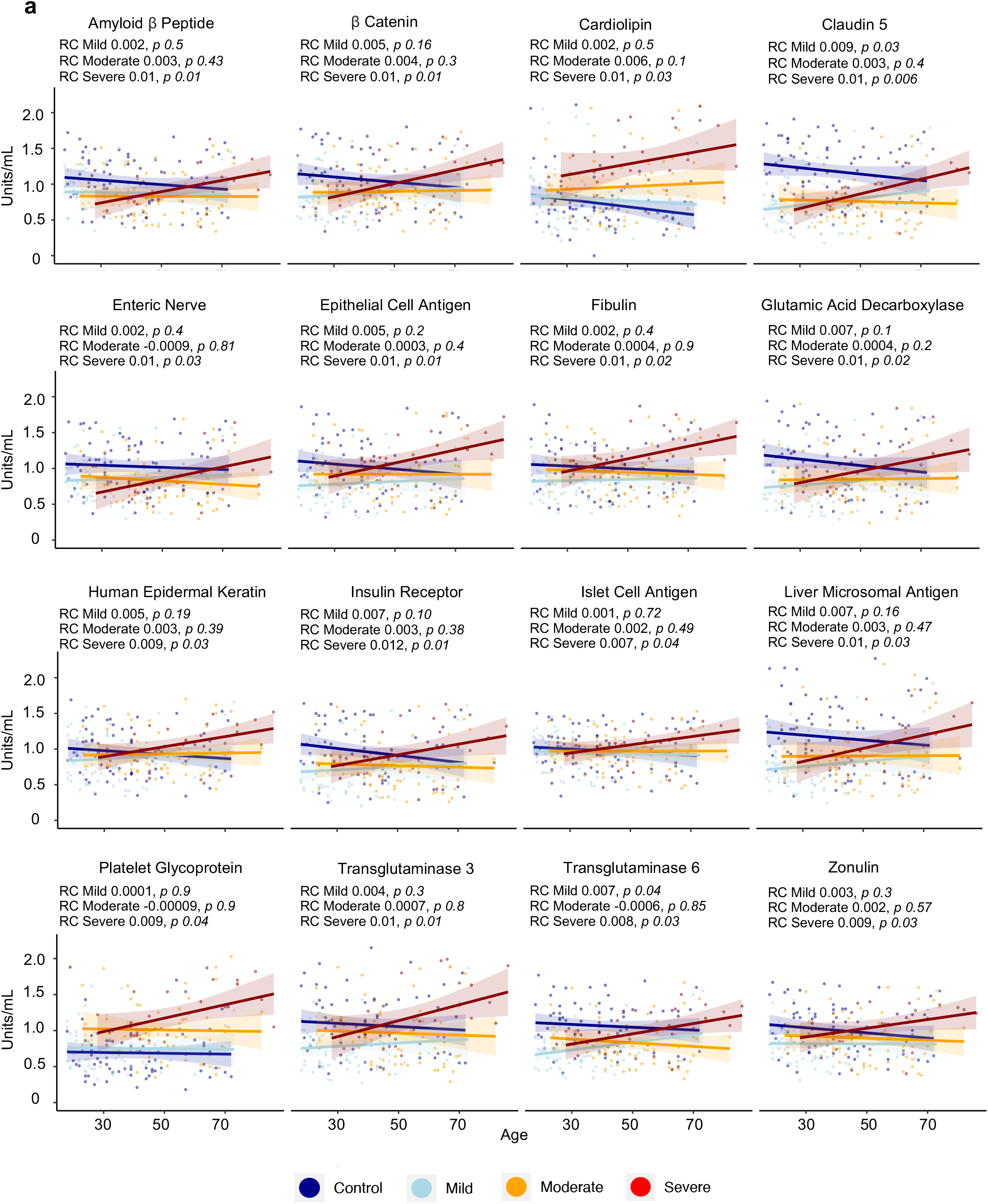
Scatter plot of regression analysis, indicating the relationship between the autoantibodies and age for healthy controls and each disease severity group according to color legend as indicated at the bottom of the graphs. The *p*-values and linear regression coefficient (RC) are shown for each group in every plot.

**Supplementary Figure 2.**
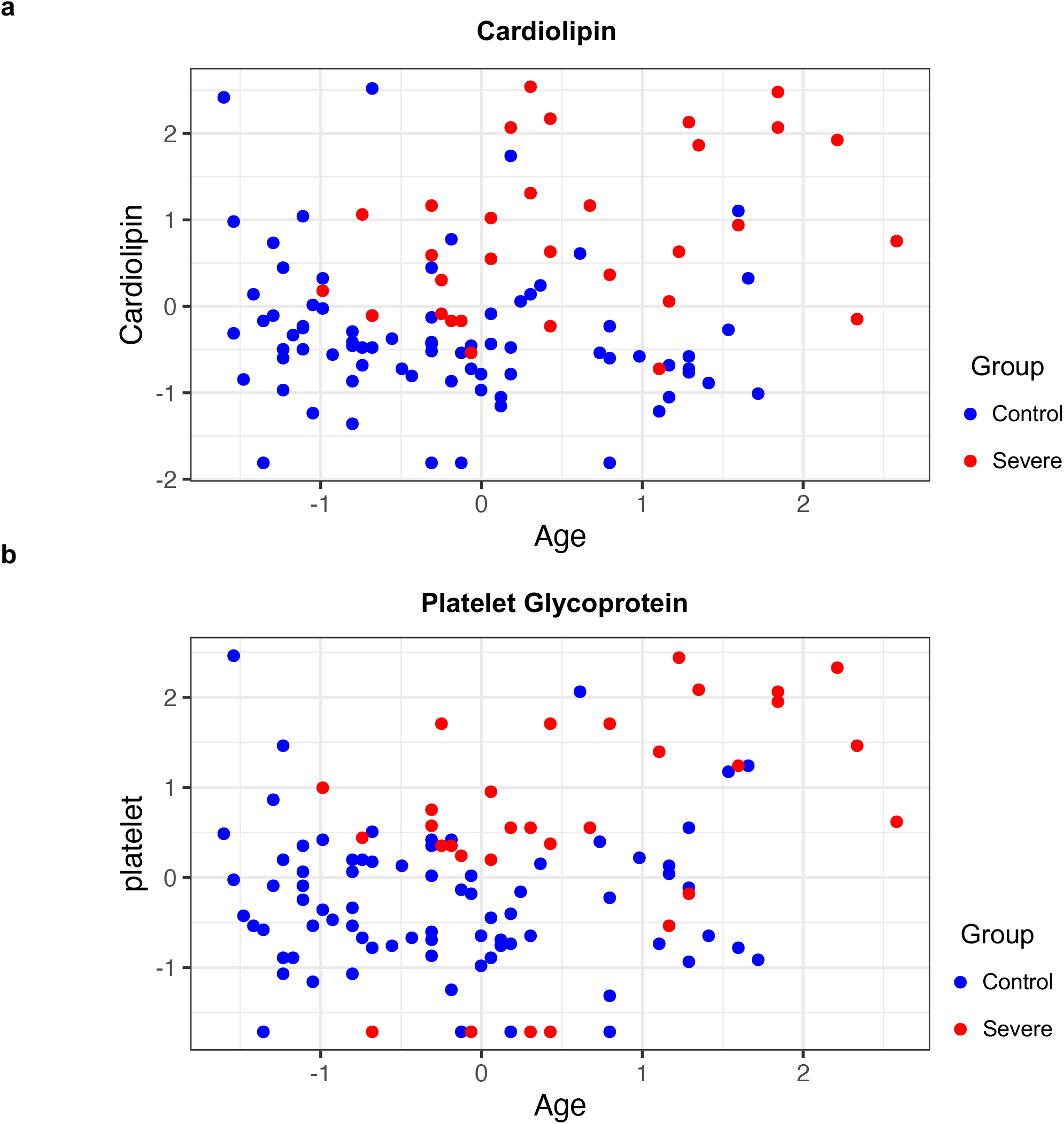
**a-b)** Scatter plots showing the distribution of elderly individuals in the healthy control (blue dots) and severe COVID-19 group (red dots) for (**a**) anti-cardiolipin antibody levels and (**b**) anti-platelet glycoprotein levels. This distribution was used as input for the SVM analysis.

**Supplementary Figure 3.**
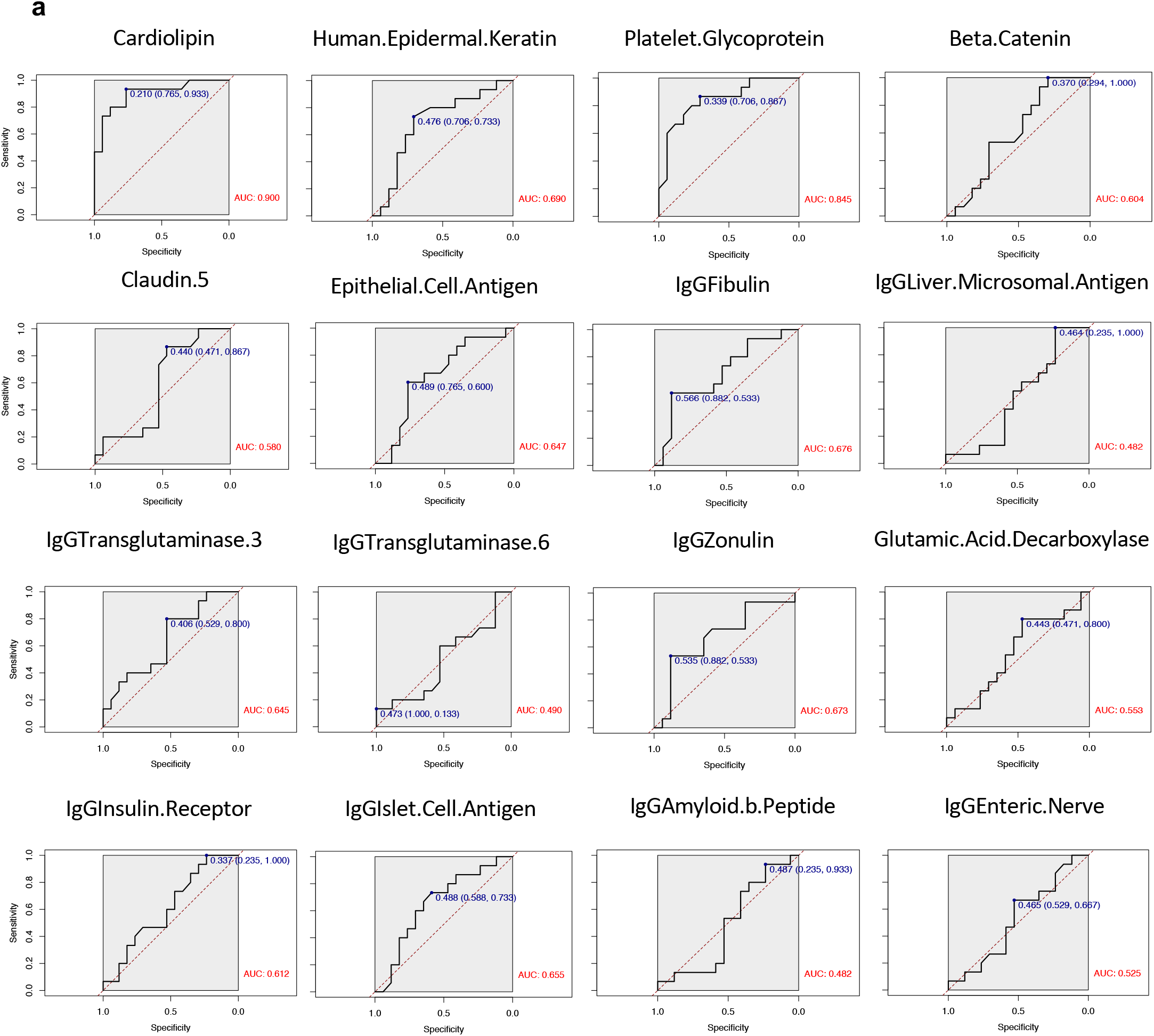
ROC curves demonstrate the specificity and sensitivity of each of the sixteen autoantibodies for severe COVID-19 in the elderly category. All plots were generated using binomial logistic regression.

## REFERENCE

1. Knight, J. S. et al. The intersection of COVID-19 and autoimmunity. J. Clin. Invest. 131, (2021).

2. Galeotti, C. & Bayry, J. Autoimmune and inflammatory diseases following COVID-19. Nat. Rev. Rheumatol. 2020 168 16, 413–414 (2020).

3. Merad, M., Blish, C. A., Sallusto, F. & Iwasaki, A. The immunology and immunopathology of COVID-19. Science (80-.). 375, 1122–1127 (2022).

4. Gregorova, M. et al. Post-acute COVID-19 associated with evidence of bystander T-cell activation and a recurring antibiotic-resistant bacterial pneumonia. Elife 9, 1–13 (2020).

5. Karami Fath, M. et al. SARS-CoV-2 Proteome Harbors Peptides Which Are Able to Trigger Autoimmunity Responses: Implications for Infection, Vaccination, and Population Coverage. Front. Immunol. 12, 3174 (2021).

6. Vojdani, A., Vojdani, E. & Kharrazian, D. Reaction of Human Monoclonal Antibodies to SARS-CoV-2 Proteins With Tissue Antigens: Implications for Autoimmune Diseases. Front. Immunol. 11, (2021).

7. Vojdani, A. & Kharrazian, D. Potential antigenic cross-reactivity between SARS-CoV-2 and human tissue with a possible link to an increase in autoimmune diseases. Clin. Immunol. 217, (2020).

8. Nunez-Castilla, J. et al. Potential Autoimmunity Resulting from Molecular Mimicry between SARS-CoV-2 Spike and Human Proteins. Viruses 14, (2022).

9. Aschman, T. et al. Association Between SARS-CoV-2 Infection and Immune-Mediated Myopathy in Patients Who Have Died. JAMA Neurol. 78, 948–960 (2021).

10. Li, Y. et al. Acute cerebrovascular disease following COVID-19: a single center, retrospective, observational study. Stroke Vasc. Neurol. 5, 279–284 (2020).

11. Moll, G. et al. MSC Therapies for COVID-19: Importance of Patient Coagulopathy, Thromboprophylaxis, Cell Product Quality and Mode of Delivery for Treatment Safety and Efficacy. Front. Immunol. 11, 1091 (2020).

12. Libby, P. & Lüscher, T. COVID-19 is, in the end, an endothelial disease. Eur. Heart J. 41, 3038–3044 (2020).

13. Rojas, M. et al. Autoimmunity is a hallmark of post-COVID syndrome. J. Transl. Med. 20, 1–5 (2022).

14. Davis, H. E. et al. Characterizing long COVID in an international cohort: 7 months of symptoms and their impact. EClinicalMedicine 38, (2021).

15. Spudich, S. & Nath, A. Nervous system consequences of COVID-19. Science (80-.). 375, 267–269 (2022).

16. Xu, E., Xie, Y. & Al-Aly, Z. Long-term neurologic outcomes of COVID-19. Nat. Med. 2022 1–10 (2022) doi:10.1038/s41591-022-02001-z.

17. Mehandru, S. & Merad, M. Pathological sequelae of long-haul COVID. Nat. Immunol. 2022 232 23, 194–202 (2022).

18. Bastard, P. et al. Autoantibodies against type I IFNs in patients with life-threatening COVID-19. Science (80-.). 370, (2020).

19. Wang, E. Y. et al. Diverse functional autoantibodies in patients with COVID-19. Nature 595, 283–288 (2021).

20. Fagyas, M. et al. The majority of severe COVID-19 patients develop anti-cardiac autoantibodies. GeroScience (2022) doi:10.1007/S11357-022-00649-6.

21. Zuo, Y. et al. Prothrombotic autoantibodies in serum from patients hospitalized with COVID-19. Sci. Transl. Med. 12, 3876 (2020).

22. Zuniga, M. et al. Autoimmunity to the Lung Protective Phospholipid-Binding Protein Annexin A2 Predicts Mortality Among Hospitalized COVID-19 Patients. medRxiv 2020.12.28.20248807 (2021) doi:10.1101/2020.12.28.20248807.

23. Losartan for Patients With COVID-19 Requiring Hospitalization - Full Text View - https://ClinicalTrials.gov.

24. Cavalli, E. et al. Entangling COVID-19 associated thrombosis into a secondary antiphospholipid antibody syndrome: Diagnostic and therapeutic perspectives (Review). Int. J. Mol. Med. 46, 903–912 (2020).

25. Dotan, A. et al. The SARS-CoV-2 as an instrumental trigger of autoimmunity. Autoimmun. Rev. 20, (2021).

26. Trahtemberg, U. et al. Anticardiolipin and other antiphospholipid antibodies in critically ill COVID-19 positive and negative patients. Ann. Rheum. Dis. 80, 1236–1240 (2021).

27. Juanes-Velasco, P. et al. SARS-CoV-2 Infection Triggers Auto-Immune Response in ARDS. Front. Immunol. 13, 1–13 (2022).

28. Chang, S. et al. New-onset IgG autoantibodies in hospitalized patients with COVID-19. Nat. Commun. 12, (2021).

29. Woodruff, M. C. et al. Relaxed peripheral tolerance drives broad de novo autoreactivity in severe COVID-19. medRxiv Prepr. Serv. Heal. Sci. (2021) doi:10.1101/2020.10.21.20216192.

30. Taeschler, P. et al. Autoantibodies in COVID-19 correlate with antiviral humoral responses and distinct immune signatures. Allergy 77, 2415–2430 (2022).

31. Baiocchi, G. C. et al. Autoantibodies linked to autoimmune diseases associate with COVID-19 outcomes Corresponding authors□: The SARS-CoV-2 infection is associated with increased levels of autoantibodies targeting immunological proteins such as cytokines and chemokines. Report. (2022).

32. Liu, Y. et al. Association between age and clinical characteristics and outcomes of COVID-19. Eur. Respir. J. 55, (2020).

33. Davies, N. G. et al. Age-dependent effects in the transmission and control of COVID-19 epidemics. Nat. Med. 2020 268 26, 1205–1211 (2020).

34. Team, F. Variation in the COVID-19 infection–fatality ratio by age, time, and geography during the pre-vaccine era: a systematic analysis. Lancet 399, 1469–1488 (2022).

35. Ma, S., Wang, C., Mao, X. & Hao, Y. R Cells dysfunction associated with aging and autoimmune disease. Front. Immunol. 10, 318 (2019).

36. López-Otín, C., Blasco, M. A., Partridge, L., Serrano, M. & Kroemer, G. The hallmarks of aging. Cell 153, 1194 (2013).

37. Chalan, P., Berg, A. van den, Kroesen, B.-J., Brouwer, L. & Boots, A. Rheumatoid Arthritis, Immunosenescence and the Hallmarks of Aging. Curr. Aging Sci. 8, 131 (2015).

38. Wang, Y. et al. Cytoplasmic DNA sensing by KU complex in aged CD4+ T cell potentiates T cell activation and aging-related autoimmune inflammation. Immunity 54, 632-647.e9 (2021).

39. Barbé-Tuana, F., Funchal, G., Schmitz, C. R. R., Maurmann, R. M. & Bauer, M. E. The interplay between immunosenescence and age-related diseases. Semin. Immunopathol. 42, 545–557 (2020).

40. Arvey, A. et al. Age-associated changes in the circulating human antibody repertoire are upregulated in autoimmunity. Immun. Ageing 17, 1–16 (2020).

41. Andrzejewska, A. et al. Multi-Parameter Analysis of Biobanked Human Bone Marrow Stromal Cells Shows Little Influence for Donor Age and Mild Comorbidities on Phenotypic and Functional Properties. Front. Immunol. 10, 2474 (2019).

42. Shome, M. et al. Serum autoantibodyome reveals that healthy individuals share common autoantibodies. Cell Rep. 39, (2022).

43. Nagele, E. P. et al. Natural IgG Autoantibodies Are Abundant and Ubiquitous in Human Sera, and Their Number Is Influenced By Age, Gender, and Disease. PLoS One 8, e60726 (2013).

44. Cabral-Marques, O. et al. Autoantibodies targeting GPCRs and RAS-related molecules associate with COVID-19 severity. doi:10.1038/s41467-022-28905-5.

45. COVID-19 Clinical management: living guidance.

46. Baiocchi, G. C. et al. Autoantibodies linked to autoimmune diseases associate with COVID-19 outcomes. medRxiv 2022.02.17.22271057 (2022) doi:10.1101/2022.02.17.22271057.

47. R: The R Project for Statistical Computing. https://www.r-project.org/.

48. RStudio | Open source & professional software for data science teams - RStudio. https://www.rstudio.com/.

49. Create Elegant Data Visualisations Using the Grammar of Graphics • ggplot2. https://ggplot2.tidyverse.org/.

50. Schneider, A., Hommel, G. & Blettner, M. Linear Regression Analysis. Dtsch. Arztebl. Int. (2010) doi:10.3238/arztebl.2010.0776.

51. Wickham, H. ggplot2: Elegant Graphics for Data Analysis. (2016).

52. A, K. rstatix: Pipe-Friendly Framework for Basic Statistical Tests. (2021).

53. Gu, Z., Eils, R. & Schlesner, M. Complex heatmaps reveal patterns and correlations in multidimensional genomic data. Bioinformatics 32, 2847–2849 (2016).

54. Gu, Z., Gu, L., Eils, R., Schlesner, M. & Brors, B. circlize Implements and enhances circular visualization in R. Bioinformatics 30, 2811–2812 (2014).

55. Ringnér, M. What is principal component analysis? Nat. Biotechnol. 2008 263 26, 303–304 (2008).

56. Lever, J., Krzywinski, M. & Altman, N. Points of Significance: Principal component analysis. Nature Methods vol. 14 641–642 (2017).

57. Sotzny, F. et al. Dysregulated autoantibodies targeting vaso- and immunoregulatory receptors in Post COVID Syndrome correlate with symptom severity. Front. Immunol. 13, 19 (2022).

58. Alboukadel Kassambara & Fabian Mundt. factoextra: Extract and Visualize the Results of Multivariate Data Analyses. (2020).

59. Liaw, A. & Wiener, M. Classification and Regression by randomForest. 2, (2002).

60. Schimke, L. F. et al. Severe COVID-19 Shares a Common Neutrophil Activation Signature with Other Acute Inflammatory States. Cells 11, (2022).

61. Kernel-Based Machine Learning Lab [R package kernlab version 0.9-31]. (2022).

62. Karatzoglou, A., Hornik, K., Smola, A. & Zeileis, A. kernlab - An S4 Package for Kernel Methods in R. J. Stat. Softw. 11, 1–20 (2004).

63. Huang, S. et al. Applications of Support Vector Machine (SVM) Learning in Cancer Genomics. Cancer Genomics - Proteomics 15, 41 LP – 51 (2018).

64. Noble, W. S. What is a support vector machine? Nat. Biotechnol. 24, 1565–1567 (2006).

65. Meyer, D., Dimitriadou, E., Hornik, K., Weingessel, A. & Leisch, F. e1071: Misc Functions of the Department of Statistics, Probability Theory Group (Formerly: E1071), TU Wien. (2021).

66. Sperandei, S. Understanding logistic regression analysis. Biochem. Medica 12–18 (2014) doi:10.11613/BM.2014.003.

67. Ricciardi, C. et al. Linear discriminant analysis and principal component analysis to predict coronary artery disease. Health Informatics J. 26, 2181–2192 (2020).

68. Venables, W. N. & Ripley, B. D. Modern Applied Statistics with S. Ripley (Springer, 2002).

69. Sachs, M. C. plotROC□: A Tool for Plotting ROC Curves. J. Stat. Softw. 79, (2017).

70. Chongsuvivatwong, V. epiDisplay: Epidemiological Data Display Package. (2022).

71. Dayimu, A. forestploter: Create Flexible Forest Plot. (2022).

72. Lever, J., Krzywinski, M. & Altman, N. Principal component analysis. Nat. Methods 14, 641–642 (2017).

73. Cabral-Marques, O. & Riemekasten, G. Functional autoantibodies targeting G protein-coupled receptors in rheumatic diseases. Nat. Rev. Rheumatol. 13, 648–656 (2017).

74. Cabral-Marques, O. et al. GPCR-specific autoantibody signatures are associated with physiological and pathological immune homeostasis. Nat. Commun. 9, 1–14 (2018).

75. Pedroza-Pacheco, I. & Borrow, P. Targeting autoantibodies in COVID-19. Nat. Rev. Immunol. 21, 134 (2021).

76. Credle, J. J. et al. Unbiased discovery of autoantibodies associated with severe COVID-19 via genome-scale self-assembled DNA-barcoded protein libraries. Nat. Biomed. Eng. 6, 992–1003 (2022).

77. Woodruff, M. C. et al. Dysregulated naive B cells and de novo autoreactivity in severe COVID-19. Nature (2022) doi:10.1038/s41586-022-05273-0.

78. Chen, Y. et al. Aging in COVID-19: Vulnerability, immunity and intervention. Ageing Res. Rev. 65, 101205 (2021).

79. Beer, J. et al. Impaired immune response drives age-dependent severity of COVID-19. J. Exp. Med. 219, e20220621 (2022).

80. Manry, J. et al. The risk of COVID-19 death is much greater and age dependent with type I IFN autoantibodies. Proc. Natl. Acad. Sci. U. S. A. 119, e2200413119 (2022).

81. Neville, C. et al. Thromboembolic risk in patients with high titre anticardiolipin and multiple antiphospholipid antibodies. Thromb. Haemost. 90, 108–115 (2003).

82. Lambert, M. P. & Gernsheimer, T. B. Clinical updates in adult immune thrombocytopenia. Blood 129, 2829–2835 (2017).

83. Katsoularis, I. et al. Risks of deep vein thrombosis, pulmonary embolism, and bleeding after covid-19: nationwide self-controlled cases series and matched cohort study. BMJ 377, (2022).

84. Quezada-Feijoo, M. et al. Elderly Population with COVID-19 and the Accuracy of Clinical Scales and D-Dimer for Pulmonary Embolism: The OCTA-COVID Study. J. Clin. Med. 10, (2021).

85. Greinacher, A. et al. Anti-platelet factor 4 antibodies causing VITT do not cross-react with SARS-CoV-2 spike protein. Blood 138, 1269–1277 (2021).

86. Nakamura, T. et al. Detection of anti-GPIbα autoantibodies in a case of immune thrombocytopenia following COVID-19 vaccination. Thrombosis research vol. 209 80– 83 (2022).

87. Liu, Q. et al. Anti-PF4 antibodies associated with disease severity in COVID-19. Proc. Natl. Acad. Sci. 119, e2213361119 (2022).

88. Fleg, J. L. & Strait, J. Age-associated changes in cardiovascular structure and function: a fertile milieu for future disease. Heart Fail. Rev. 17, 545–554 (2012).

89. Thomas, E. T., Guppy, M., Straus, S. E., Bell, K. J. L. & Glasziou, P. Rate of normal lung function decline in ageing adults: a systematic review of prospective cohort studies. BMJ Open 9, e028150 (2019).

90. de Almeida Chuffa, L. G. et al. Aging whole blood transcriptome reveals candidate genes for SARS-CoV-2-related vascular and immune alterations. J. Mol. Med. 100, 285–301 (2022).

91. Páez-Franco, J. C. et al. Metabolomics analysis identifies glutamic acid and cystine imbalances in COVID-19 patients without comorbid conditions. Implications on redox homeostasis and COVID-19 pathophysiology. PLoS One 17, 1–17 (2022).

92. Ludwig, R. J. et al. Mechanisms of Autoantibody-Induced Pathology. Front. Immunol. 8, 603 (2017).

93. Pacheco, Y. et al. Cytokine and autoantibody clusters interaction in systemic lupus erythematosus. J. Transl. Med. 15, 239 (2017).

94. Grolleau-Julius, A., Ray, D. & Yung, R. L. The role of epigenetics in aging and autoimmunity. Clin. Rev. Allergy Immunol. 39, 42–50 (2010).

95. Cao, X. et al. Accelerated biological aging in COVID-19 patients. Nat. Commun. 13, 2135 (2022).

96. Farris, A. D. & Guthridge, J. M. Overlapping B cell pathways in severe COVID-19 and lupus. Nat. Immunol. 21, 1478–1480 (2020).

97. Bertin, D. et al. Anti-cardiolipin IgG autoantibodies associate with circulating extracellular DNA in severe COVID-19. Sci. Rep. 12, 12523 (2022).

98. Manoussakis, M. N. et al. High prevalence of anti-cardiolipin and other autoantibodies in a healthy elderly population. Clin. Exp. Immunol. 69, 557–565 (1987).

99. Njemini, R. et al. The prevalence of autoantibodies in an elderly sub-Saharan African population. Clin. Exp. Immunol. 127, 99–106 (2002).

100. Tripathi, U. et al. SARS-CoV-2 causes senescence in human cells and exacerbates the senescence-associated secretory phenotype through TLR-3. Aging (Albany. NY). 13, 21838–21854 (2021).

101. Bartleson, J. M. et al. SARS-CoV-2, COVID-19 and the aging immune system. Nat. Aging 1, 769–782 (2021).

102. Collier, D. A. et al. Age-related immune response heterogeneity to SARS-CoV-2 vaccine BNT162b2. Nature 596, 417–422 (2021).

103. Lynch, S. M., Guo, G., Gibson, D. S., Bjourson, A. J. & Rai, T. S. Role of Senescence and Aging in SARS-CoV-2 Infection and COVID-19 Disease. Cells 10, (2021).

104. Akbar, A. N. & Gilroy, D. W. Aging immunity may exacerbate COVID-19. Science (80-.). 369, 256–257 (2020).

105. Hou, Y. et al. Aging-related cell type-specific pathophysiologic immune responses that exacerbate disease severity in aged COVID-19 patients. Aging Cell 21, e13544 (2022).

